# Identification of Nonverbal Communication Tools for Use in Dementia

**DOI:** 10.1101/2021.01.28.21250369

**Authors:** Zunera Khan, Miguel Vasconcelos Da Silva, Jemuwem Eno-Amooquaye, Steven Nowicki, Kayleigh Nunez, Paul Francis, Clive Ballard

## Abstract

**Introduction:** Non-verbal communication remains a relatively unexplored area in dementia care with a lack of validated assessment tools available to measure non-verbal communication function in dementia.

**Methods:** This scoping review identifies assessment scales of nonverbal communication in dementia and evaluates the psychometric properties and clinical utility of these instruments. Relevant publications in English, from 1947 to 2017, were identified through an extensive search strategy in Medline, Psychinfo and the Cumulative Index to Nursing and Allied Health Literature (CINAHL), EMBASE, Cochrane and generic search engines (Google) and available off-line resources. Quality judgement criteria was formulated and used to evaluate the psychometric aspects of the scales.

**Results:** Forty-one tools were identified measuring various communication channels including verbal, nonverbal (e.g., facial expressions, gestures, eye contact) and functional, communication means; within various settings and populations, for instance, those assessing cognition and verbal language difficulties secondary to stroke, aphasia and nonverbal cues associated with pain. A number of tools presented psychometrics qualities; only nine of the forty-one tools specifically focussed on nonverbal communication, however, comprehensive assessment of nonverbal communication function was not presented in majority of the identified tools. Two tools provided a detailed assessment of nonverbal communication, the Emory dyssemia Index (EDI) and the Threadgold Communication Tool (TCT).

**Conclusion:** Based on the psychometric qualities and criteria regarding sensitivity and clinical utility, we concluded that although it is difficult to recommend one particular tool, the EDI and TCT are the most appropriate scales currently available. Further research should focus on improving these scales by further testing their validity, reliability and clinical utility in dementia.

## Background

Communication impairment may interrupt various aspects of daily living for people living with Alzheimer’s Disease (PlwD) and may be associated with changes in behaviour or distress, such as agitation, hostility and physical aggression, usually reflecting an attempt to communicate [1, 2]. The use of outcome measures has long been held as a gold standard in research and they often undergo a rigorous development procedure [3]. Several tools have been developed to assess different aspects of communication for PlwD, most of which focus on the person’s expression in terms of agitation and aggression rather than the ability to communicate [4]. Limited research literature is present on assessing nonverbal communication and to date no overview is available of communication scales developed specifically to assess nonverbal communication in PlwD. There is thus a need to identify manageable, valid and reliable tools for assessing nonverbal communication function/impairment in PlwD. The ideology was to find an assessment tool that could be used readily in daily practice and to assess an effectiveness of an intervention with pre- and post-assessment of nonverbal communication function. This review aimed to identify measures assessing communication function, specifically nonverbal communication and conduct the appraisal of these measures’ psychometric properties using data from earlier studies to guide future choice of measures in research and practice within dementia.

## Summary of key Objectives

This paper aims to identify feasible research tools for measuring communication function in PlwAD specifically nonverbal communication. Two research questions were addressed to:

- Identify nonverbal communication assessment tools that are available to assess nonverbal communication function in the current literature.
- Assess the psychometric qualities of the identified tools, including validity measures containing content validity, criterion validity and construct validity, test re-test reliability and interrater reliability.

## Design

Scoping review of the literature to identify outcome measures used within research for assessing nonverbal communication among people living with dementia.

## Methods

The literature search was conducted through electronic databases and hand searches. The review process utilised publications based on an extensive search strategy, involving database searches of Medline, Psychinfo and the Cumulative Index to Nursing and Allied Health Literature (CINAHL), EMBASE, Cochrane and generic search engines (Google) and available off-line resources Two forms of hand-searches were undertaken: the references and bibliographies of key articles retrieved in the literature search from above search engines. The title of papers published from 1946 to 2017 were scrutinised.

### Rationale for the scoping review

A scoping review was conducted following established methods [5-8]. Scoping studies can be particularly relevant to disciplines with emerging evidence, in which the paucity of literature makes it difficult for researchers to undertake systematic reviews. Arksey and O’Malley (6) defined a framework of how to conduct a scoping review and outlined a scoping review as a process which focuses on the appraisal of a body of studies examining the extent range, and nature of existing literature.

A scoping review was chosen for the current study because this methodology aligned with the broad research objective and allowed for the inclusion of research papers, studying body of literature to identify research tools available for assessment of nonverbal cues. Utilisation of the scoping review methodology allowed for the collection of literature, which provides an overview of our current understanding of nonverbal communication tools within dementia, AD and learning disabilities (LD) within the research field. Furthermore, by following the established methods as dictated by the scoping review framework enabled the systematic and comprehensive appraisal of literature.

### Search strategy and inclusion criteria

The literature search strategy was structured using the relevant concepts of PICO (Population, Intervention, Comparator, Outcome). Setting was not controlled for the purpose of the literature search with the aim to find broad range of tools assessing nonverbal communication across various settings.

The following search terms were used when running searches through above listed electronic databases:

- nonverbal communication assessment
- nonverbal communication tools
- the nonverbal communication assessment tool
- nonverbal communication questionnaire
- nonverbal communication instrument
- nonverbal communication scale *’nonverbal’ was also entered as well as ‘non-verbal’
- *’Alzheimer’s Disease’ and ‘aphasia’ and ‘dementia’ and learning disabilities (LD) were added to each of these search terms and the search run again to cover significant supplementary literature in relation to availability of nonverbal communication scales. Limited literature within Alzheimer’s Disease and dementia was expected thus the literature search was expanded to include other parallel areas and terms that were likely to contribute to additional information, such as LD and aphasia. Disease severity or form of dementia was not specified to gauge a broader scope of literature covering nonverbal communication.

#### Inclusion criteria

1. Inclusion of a tool assessing elements of communication including nonverbal cues within Alzheimer’s disease, Dementia, Aphasia, Learning Disabilities.
2. Inclusion of a tool which can be proxy-reported, self-reported or the use of Test Battery
3. Availability of the full text in English.

#### Exclusion criteria

1. No reporting of communication measures.
2. Studies published in a language other than English if a translation was not available.
3. Development information for outcome measures was not freely available.
4. Case reports or secondary sources/reviews

### Electronic Database Search

Electronic databases were filtered for English only articles, following that, duplicated articles were also filtered out. Initial review of the titles was conducted to extract articles which signposted elements of communication (both verbal and nonverbal). Review of titles was followed by a review of abstracts, which included both verbal and nonverbal communication elements. Papers with full-text availability were reviewed to extract information for individual tools.

### Expert consultation

In addition to the above literature search process through relevant databases, an expert consultation was also conducted to ensure coverage of communication tools examining nonverbal communication function. Research experts, who were highlighted through references of key articles, were contacted via an email by the researcher. Five key researchers; Professor Nowicki Steven, Professor Valerie Manusov, Dr Judith Saxton, Dr Carol Magai and Dr Sheila Wirz were contacted for this purpose. These included researchers who had published key papers focusing on communication in addition to cognitive impairment. It was felt that breadth and depth of knowledge of this group regarding the area of communication would provide sufficient coverage to identify any papers or assessment being utilised to examine nonverbal communication that had not been identified.

Tools identified through the electronic databases, hand searches and the expert consultation were further taken into the next stage for data extraction and quality assessment (Figure 11).

**Figure 1.**
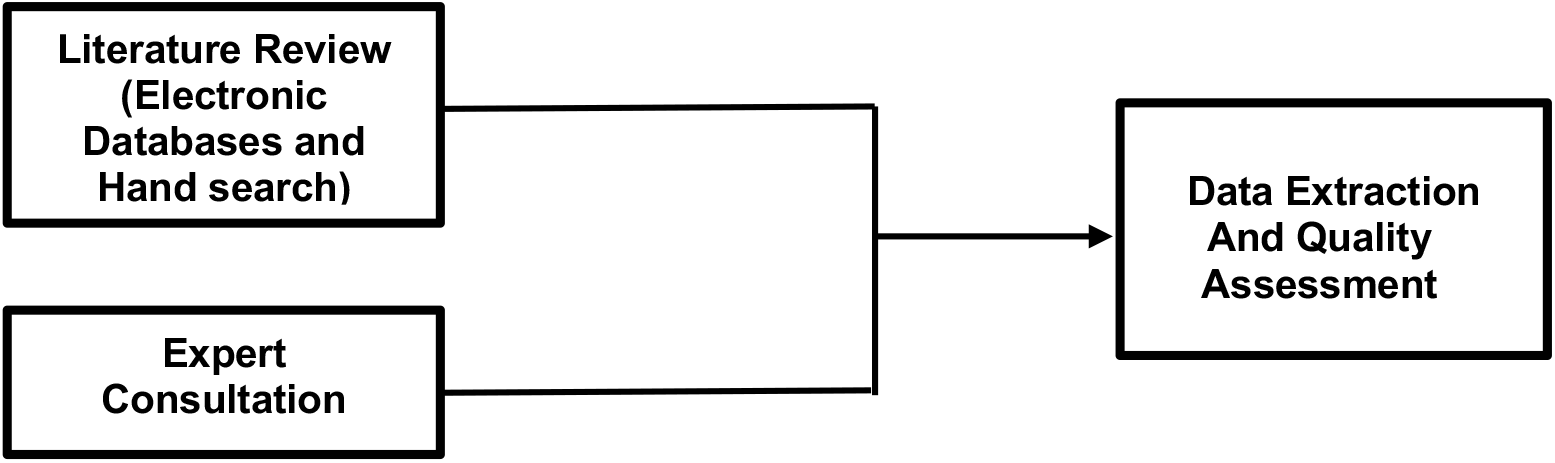
Literature Review Stages.

### Data Extraction and Quality Assessment

Full texts of articles for all of the tools were identified through electronic databases, hand search and expert consultation. Reviewer discrepancy was checked, discussed and moderated by a third reviewer. The data abstraction criteria were partly based on Streiner and Norman’s requirements for health measurement scales for the evaluation of identified tools [9]. Pairs of reviewers independently screened titles and abstracts. The following data were extracted (if available) to examine the nature and methodological quality of the assessment scales: the tool name, publication year, type of tool target population, the source of the items (origin), sample size, psychometric properties and methodology including feasibility and time taken to complete. Psychometric qualities were based on validity measures containing content validity, criterion validity and construct validity in addition to test re-test reliability and interrater reliability. Following the Streiner and Norman’s criteria, individual tool was rated against each individual extracted data item and sum for psychometrics quality was generated. In addition, an assessment for suitability was also conducted on each tool, examining assessment coverage of nonverbal communication function, i.e., a person’s ability to express and receive nonverbal cues.

**Figure 2.**
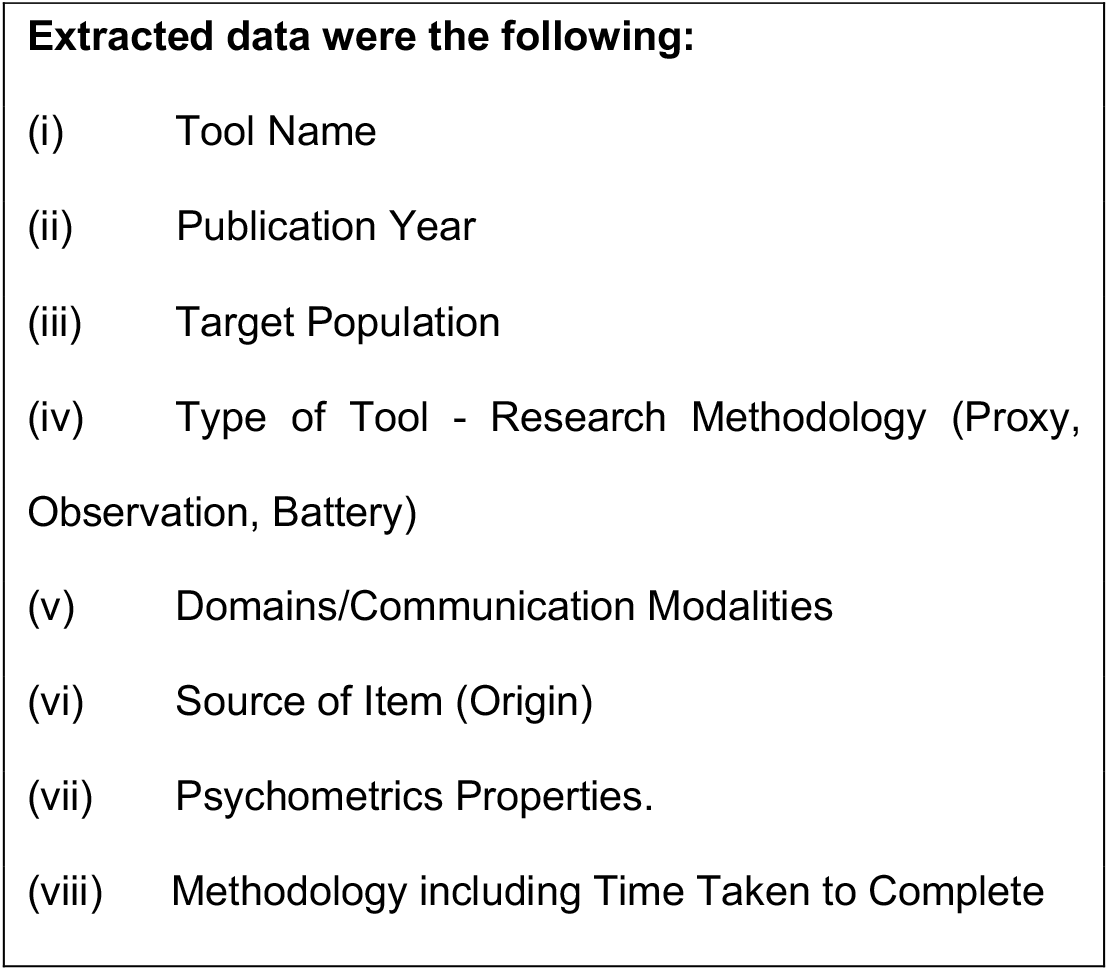
Extracted Data for each Communication Tool.

### Extended literature review of papers published from 2017 to 2019

A narrative literature review was conducted to extend the findings of the scoping review (2017). The aim was to identify any supplementary nonverbal communication tools to date (Oct 2019) post the initial scoping review.

## Methods

Searches across EMBASE, MEDLINE and Google scholar were performed for articles published from June 2017 to Oct 2019 using the following key terms: This literature may not fulfil the inclusion and exclusion criteria for the scoping review but were reported in the current chapter because they were likely to represent supplementary literature.

nonverbal communication tools

- nonverbal communication assessment tool
- nonverbal communication scale
- *’nonverbal’ was also entered as well as ‘non-verbal’
- *’Alzheimer’s’ and ‘dementia and aphasia.

## Results

A total of 1183 titles were identified through database searches, of which 939 titles were excluded based on lack of reference to verbal and nonverbal communication in the title of the paper. In total, 244 titles were screened for the inclusion of papers, 114 abstracts signposted components of both verbal and nonverbal communication and abstracts, covered elements of nonverbal communication and verbal communication tools. The final literature search review included 36 papers, which were obtained from database searches, and provided full-text information on the methodology used. In addition, five tools were identified through consultation with experts and hand searching from the reference lists of the papers that were already part of the review process. Hand searches led to three tools being identified and two being identified through expert consultation.

**Figure 3.**
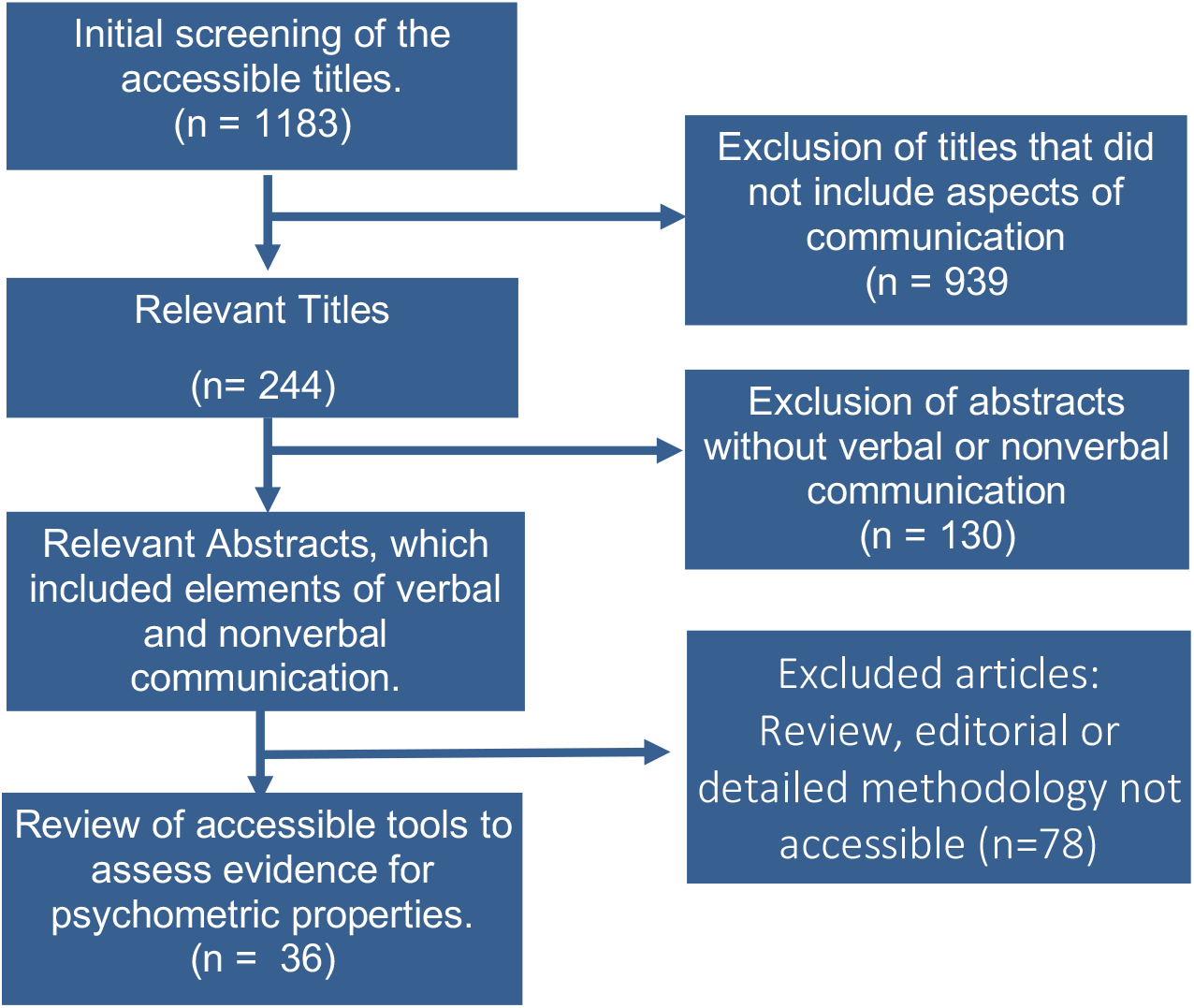
PRISMA flowchart of Literature Search Process through Electronic Databases.

### Expert Consultation

Of the five researchers contacted via an email, four responded. Tools including the Emory Dyssemia Tools (EDI) [10], American Speech Language Hearing Association Functional Assessment Of Communication Skills For Adults (ASHA-FACS) [11], were identified through this process.

### Data Extraction

#### Types of Tools and Settings

The majority of identified tools (28/41) were observational, conducted by healthcare professionals or trained other professionals. Other tools included neuropsychological batteries for testing aphasia (12/41) with one structured interview.

The tools were assessed in a range of settings and with various populations. Identified tools were examined in targeted population including survivors of paediatric brain tumours, students, individuals with aphasia, people with developmental disorders, PlwD, adults, older adults, nursing home residents, inpatients in ICU, patients with traumatic brain injury, people with stroke and people with autism.

The list of settings where these tools were used included: Class (school), high school, neuro-rehabilitation units, hospital, inpatient dementia special care units, long-term care, long-term care facilities, intensive care unit, care homes, residential aged care homes, nursing homes, independent practice, managed care and academic, clinic, palliative care hospitals, critical care units, aphasic patients’ homes, inpatient rehabilitation unit, speech therapy department, stroke unit, rehabilitation department, community, domus units in long-stay mental hospitals, psychiatric facilities, homes for the aged, continuing care hospitals and residential care facilities.

Some of these tools were required to have video recording, proxy questionnaires, checklists with live and static observations. Other methods involved the use of batteries such as the Diagnostic Analysis of Nonverbal Accuracy-2 (DANVA-2), Emotion Recognition Task (ERT) and Posture knowledge test to assess specific elements of nonverbal communication. Since the review was quite broad, with the inclusion of both verbal and nonverbal communication, some of the tools associated with stroke and aphasia purely focussed on verbal language batteries.

### Psychometric Quality Assessment

Information on psychometric elements such as content, criterion and construct validity inter-rater reliability and re-test reliability are presented below for individual tools that were found as part of the review process.

### Reliability

Seven tools showed limited evidence for reliability (REFCP, STALD, BDAE, Core Linguistic Battery, Posture Knowledge Test, Social Competence Scale and FLCI). The sections below provide further detail on three different elements of reliability.

### Interrater reliability

Data extracted for measuring inter-rater reliability using intra-class correlations were recorded as following: ACIS (0.92), ASHA FACS (0.998) Abbey pain scale (0.44-0.63), CETI (0.73), DS-DAT (0.97), PACSLAC (0.77-0.96), Doloplus-2 (0.77), MOSES (0.58 − 0.97), PADE (0.54-0.96). Kappa coefficients were provided for the FLACC (0.404), CNPI (0.63-0.82), QUIS (0.60 − 0.91), NOPAIN (0.87), LCT (0.90) and CPOT (k = 0.52 − 0.88). Correlation coefficients were used to assess agreement for; FACS (0.82-0.92), NVPS (0.80–0.87) and PAINAD (0.72-0.97). In addition, percentage agreement was used to calculate agreement for the FAST with 93% agreement between raters.

No inter-rater reliability data was presented for seventeen tools (EDI, BDAE, STALD, DANVA-2, PICA, CADL, Holden Communication Scale, KNVECL, Core Linguistic Battery Communication Problems Scale, ERT, Posture Knowledge Test, Social Competence Scale, FLCI, CASL, Verbal Fluency FAS, Interpersonal Communication Inventory and the Adult Behaviour Questionnaire).

Overall, reliability measures were carried out on small sample size and raters, so data for all of the tools are limited, thus restricting the generalisability of findings.

### Test-retest Reliability

Examination of intra-rater or Test-retest reliability involved percentage agreement, correlation, kappa, and intra-class correlations. Data extracted for examining intra-rater reliability using intra-class correlations reported the following: PADE (0.70-0.98), PACSLAC (0.86), CETI (0.94), ASHA FACS (0.995). In addition, correlation coefficient between test and retest showed the following findings: DS DAT (0.60), NVPS (0.51 − 0.75), PICA (0.96), CADL-2 (0.903), Verbal Fluency FAS (0.74), Interpersonal Communication Inventory (0.86), CASL (0.92-0.93), ERT (0.53-0.71) and FACS (0.88-0.97). Kappa coefficients were calculated for FAST with reliability was found to be excellent (kappa = 1.00), having a perfect agreement between the two test administration times. However, there were no test-retest reliability data reported for ADD, Doloplus-2, REFCP, STALD, PAINAD, FLACC, Abbey Pain Scale, CNPI, NDEPT, CPOT, BDAE, BPS, KNVECL, Lille Communication Test, QUIS, ACIS, Core Linguistic Battery, Communication Problems Scale, MOSES, NOPPAIN Posture Knowledge Test, Social Competence Scale, FLCI, FACS and ABQ.

### Internal consistency

Internal consistency was reported for the following CNPI (0.54-0.64), Doloplus-2 (0.668-0.82), PACSLAC (0.74-0.92), PADE (0.24-0.88), PAINAD (0.5-0.74), EDI (α = 0.97), DANVA-2 (α = 0.64-0.77), PAINAD (α = 0.5-0.65), FLACC (α = 0.882), Abbey Pain Scale (α = 0.74–0.81), DS DAT (α = 0.77), CPOT (α = 0.81), PADE (α = 0.77–0.88), PACSLAC (α = 0.82–0.86), KNVECL (α = 0.803), Doloplus-2 (α = 0.668-0.82), NVPS (α = 0.76), CETI (α = 0.94), CADL-2 (α = 0.64 −0 .94), HCS (α = 0.94), Communication Problems Scale (α = 0.88), MOSES (α = 0.79 − 0.82), CASL (α = 0.85 − 0.96), ABQ (α = 0.48), ASHA FACS (α = 0.95) and FAS with the coefficient alpha of r = .83 was sufficiently high to ensure high item homogeneity even though t-tests showed the number of words was significantly different (α = .001) among the three letters (F vs. A: t(1,893) 5 19.0; F vs. S: t(1,893) 5 5.2; A vs. S: t(1,893) 5 23.0). Contrarily, there were a number of tools for which data relating to internal consistency was not available. These included the REFCP, STALD, NDEPT, NOPPAIN, FAST, PICA, ADD, LCT, QUIS, ACIS, FLCI, Core Linguistic Battery, ERT, Posture Knowledge Test, Social Competence Scale, FACS and Interpersonal Communication Inventory.

### Validity

The validity of the identified tools was examined using content, construct and criterion validity, these are further detailed in the subsequent sections. Twelve of the 41 tools seemed to have limited evidence for validity assessment. These included: REFCP, NDEPT, NOPPAIN, CADL-2, KNVECL, ADD, LCT, Core Linguistic Battery, Posture Knowledge Test, Social Competence Scale, FLCI and Interpersonal Communication Inventory.

### Criterion validity

Criterion validity was evaluated by examining the correlation of scores between tools. Overall, the tools with the highest correlations included EDI correlation with Behavioural Rating Scale (r = − 0.74), STALD with Frenchay Aphasia Screening Test (r = 0.74), PAIN AD with DS DAT (r = 0.76), FLACC with CNPI (r = 0.96), Frenchay Aphasia Screening test with Functional Communication Profile (r = 0.73), PICA with CET (r = 0.63) and western aphasia battery, ASHA FACS with ADAS Cog (r = 0.63) and with MMSE r = 0.72), HCS with MMSE (r = − 0.67) and CASL with Peabody picture vocabulary (r =0.74). Data was not available for review for 18 tools. Other small to moderate associations were reported between DANVA-2 and PONS (r = 0.44), CNPI and VDS (r =.372), self-report VAS (r = − 0.30-0.50), CPOT and patients’ self-report ratings (r = 0.49), PADE and CMAI (r = 0.30 − 0.42), PACSLAC and Doloplus 2 (r=0.34), DS DAT and Pittsburgh Agitation Scale (r = 0.51), ERT and Interpersonal Behaviour Checklist (r = 0.53), MOSES Depression Anxiety and Zung Depression Status Inventory (R = 0.49), ABQ Joy and facial expressions (r= 0.61).

### Construct validity

Construct validity was measured by comparing scores to function and disease severity. Fifteen tools did not exhibit data in relation to construct validity. Table 5 provides a summary of construct validity across tools. Tools including PACSLAC, CETI, Frenchay Aphasia Screening Test, PICA, ASHA FACS, ERT and MOSES scored high for construct validity.

**Table 1.** Inclusion / Exclusion Criteria for the literature search.

| Parameter | Inclusion | Exclusion |
| --- | --- | --- |
| Population | Alzheimer's disease, Dementia, Aphasia, Learning Disabilities | Not applied |
| Intervention | Communication Tools | No reporting of communication tools |
| Comparator | Not Applied | Not Applied |
| Outcome | Nonverbal, verbal communication, functional communication | No reporting of communication measures |

**Table 2.**
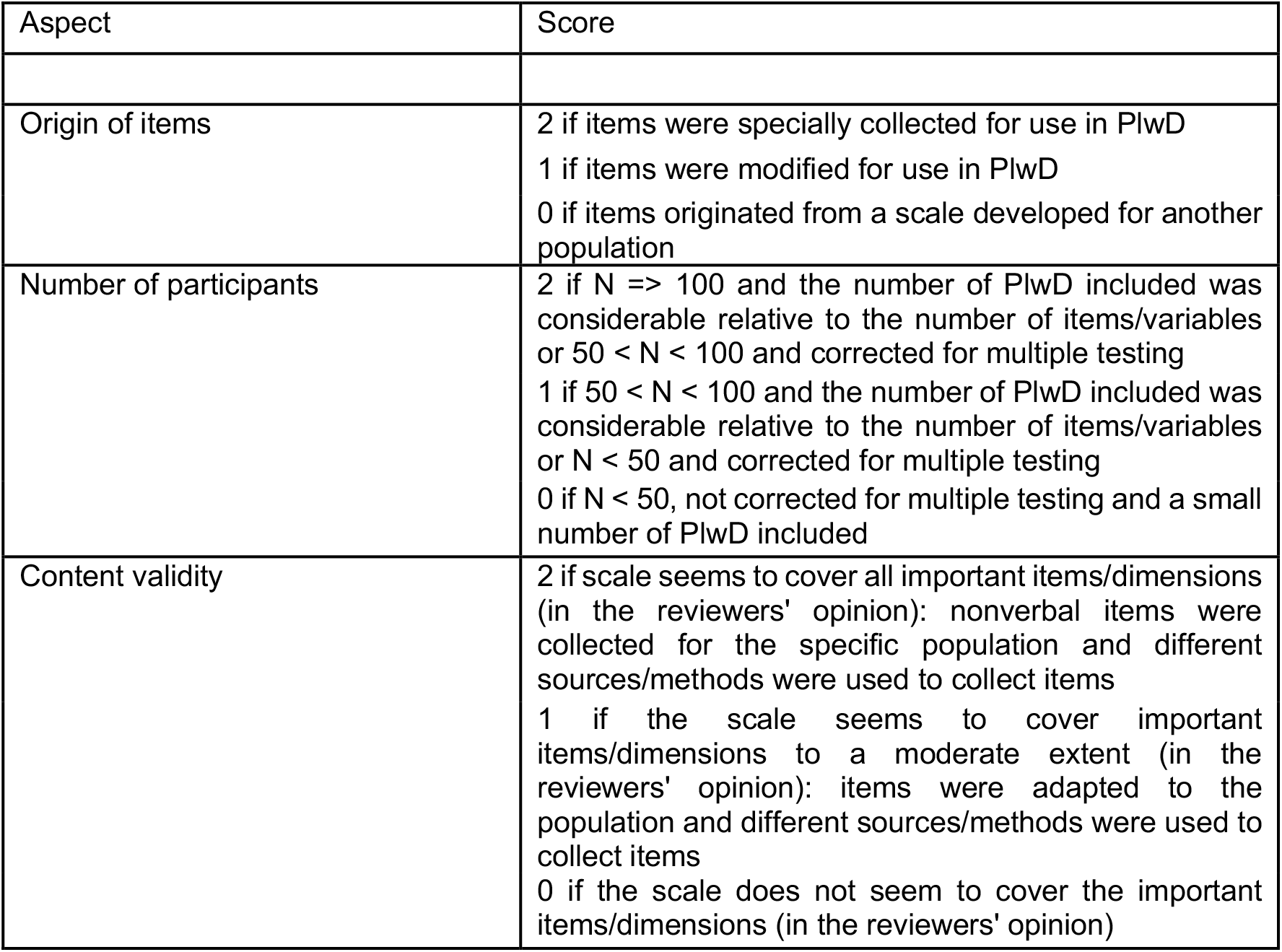
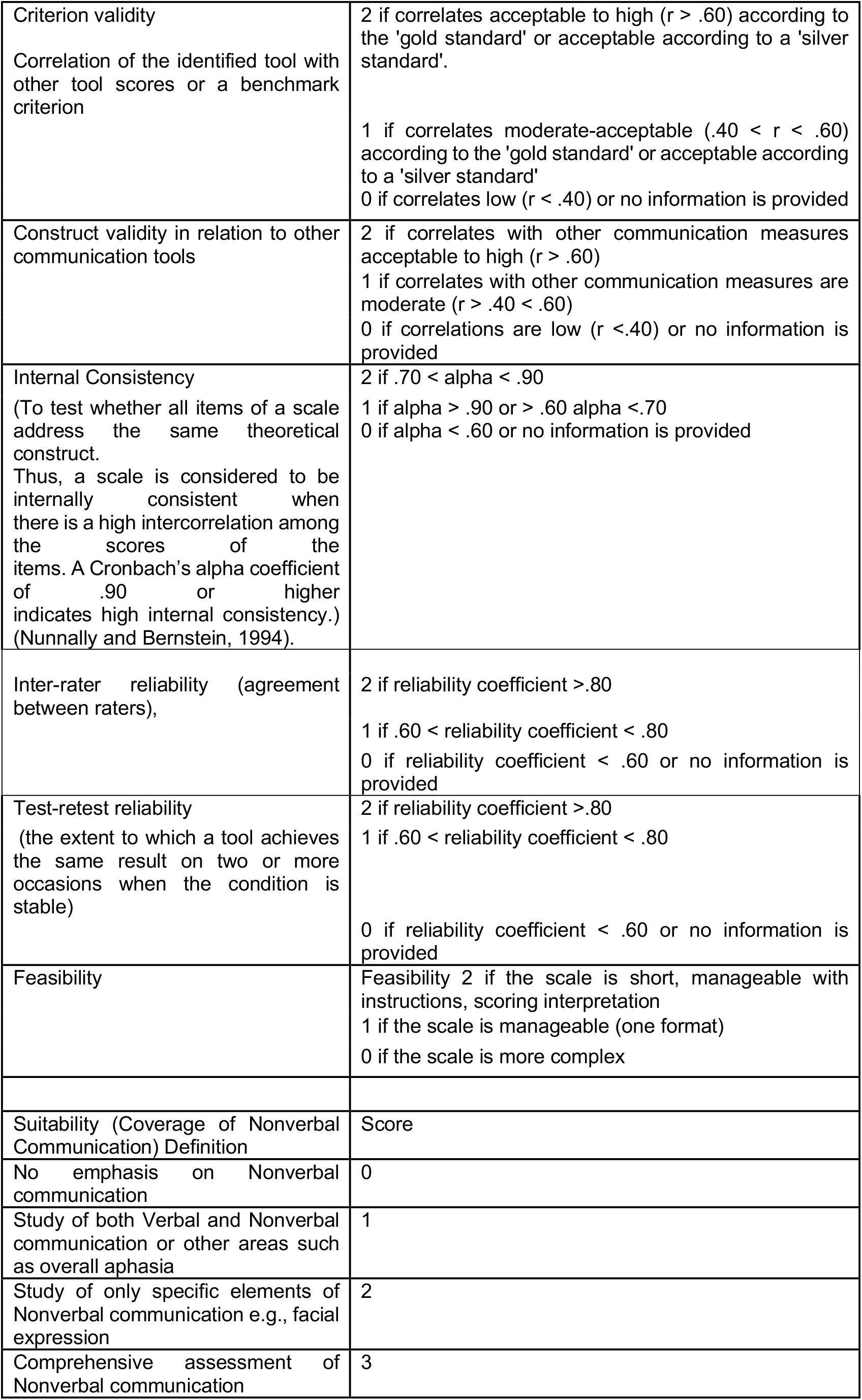
Data Abstraction Criteria based on Psychometric properties.

**Table 3.** Assessment Tools and their modalities.

| **Assessment Tool** | **Modality** |
| --- | --- |
| Emory Dyssemia Index (EDI) [10] | Nonverbal Communication |
| Revised Edinburgh Functional Communication Profile (REFCP) [12] | Functional Communication |
| Sheffield Screening Test for Acquired Language Disorders (STALS) [13] | Verbal communication |
| Diagnostic Analysis of Nonverbal Accuracy 2 (DANVA-2) [14] | Nonverbal Communication |
| Pain Assessment in Advanced Dementia (PAINAD) [15] | Pain |
| Face, Legs, Activity, Cry, Consolability) Pain Scale (FLACC) [16] | Pain |
| Abbey Pain Scale [17] | Pain |
| Checklist of Nonverbal Pain Indicators (CNPI) [18] | Pain |
| Nonverbal communication in doctor–elderly patient transactions (NDEPT) [19] | Nonverbal communication |
| Discomfort Scale - Dementia for Alzheimer Type (DS DAT) [20] | Pain |
| Critical Care Pain Observation Tool (CPOT) [21] | Pain |
| Pain Assessment for the Dementing Elderly (PADE) [22] | Pain |
| Pain Assessment Checklist for Seniors with Limited Ability to Communicate) (PACSLAC) [23] | Pain |
| Behavioral Pain Scale (BPS) [24] | Pain |
| Doloplus-2 [25] | Pain |
| NOPPAIN (Non-Communicative Patient's Assessment Instrument [26] | Pain |
| NVPS: Nonverbal Pain Scale [27] | Pain |
| Communication Effectiveness Index (CETI) [28] | Functional Communication |
| Boston Diagnostic Aphasia Examination (BDAE) [29] | Aphasia |
| The Frenchay Aphasia Screening Test [30] | Aphasia |
| Porch Index of Communicative Abilities (PICA) [31] | Functional Communication |
| Communication Activities of Daily Living -2 (CADL-2) [32] | Functional Communication |
| ASHA Functional Assessment of Communication Skills (ASHA FACS) [11] | Functional Communication |
| Kovacev Non-Verbal Expression Check List (KNVECL) [33] | Nonverbal Communication |
| ADD (Assessment of Discomfort in Dementia) [34] | Pain |
| Lille Communication Test (LCT) [35] | Verbal and Nonverbal Communication |
| The Holden Communication Scale (HCS) [36] | Verbal and Nonverbal Communication |
| Quality of Interaction schedule (QUIS) [37] | Verbal and Nonverbal Communication |
| The Assessment of Communication and Interaction Skills (ACIS) [38] | Verbal and Nonverbal Communication |
| Core Linguistic Battery [39] | Overall communication |
| Communication Problems Scale [40] | Verbal Communication Problems |
| Emotion Recognition Test (ERT) [41] | Nonverbal communication |
| Multidimensional Observation Scale for Elderly Subjects (MOSES) [42] | Multimodal functions |
| Posture Knowledge Test [43] | Nonverbal Communication |
| Social Competence Scale [44] | Social Competence |
| The Functional Linguistic Communication (FLCI) Inventory [45] | Functional Communication |
| Comprehensive Assessment of Spoken Language (CASL) [46] | Verbal communication |
| Facial Action Coding System (FACS) [47] | Nonverbal communication |
| Verbal Fluency Benton (FAS) [48] | Aphasia |
| Interpersonal Communication Inventory [49] | Verbal and Nonverbal Communication |
| The Adult Behavior Questionnaire (ABQ) [50] | Nonverbal |

**Table 4.** Criterion Validity of Tools identified.

| Assessment Tool | Criterion (correlation with other tools) |
| --- | --- |
| Emory Dyssemia Index (EDI) (Nowicki and Duke, 1992) | $r = -0.74$ with Behavioral Rating Scale (BES-2; Mann & Kenowitz, 1985) |
| Revised Edinburgh Functional Communication Profile (REFCP) (Wirz and Skinner, 1990) | Not Available |
| Sheffield Screening Test for Acquired Language Disorders (STALD) (Syder, 1993) | $r = 0.74$ with Frenchay Aphasia Screening Test |
| Diagnostic Analysis of Nonverbal Accuracy 2 (DANVA-2) (Duke and Nowicki, 1993) | $r = 0.44$ Profile of Nonverbal Sensitivity (PONS) |
| Pain Assessment in Advanced Dementia (PAINAD) (Warden et al., 2003) | $r = 0.76$ with DS DAT, $r = 0.75$ with Pain VAS |
| Face, Legs, Activity, Cry, Consolability) Pain Scale (FLACC) (Willis et al., 2003) | $r = 0.96$ with CNPI (Checklist of Nonverbal Pain Indicators) (Feldt, 2000) |
| Abbey Pain Scale (Abbey et al., 2004) | Not Available |
| Checklist of Nonverbal Pain Indicators (CNPI) (Feldt, 2000) | r = .372 with Verbal Descriptor Scale (VDS), r = - 0.30-0.50 with Self report VAS |
| Nonverbal communication in doctor–elderly patient transactions (NDEPT) (Gorawara-Bhat, 2014) | Not Available |
| Discomfort Scale - Dementia for Alzheimer Type (DS DAT) (Hurley et al., 1992) | r=0.76 with PAINAD, r = 0.51 with Pittsburgh Agitation Scale |
| Critical Care Pain Observation Tool (CPOT) (Gelinas et al., 2006) | r = 0.49, between patients' self-reports of pain and the scores on the Critical-Care Pain Observation Tool |
| Pain Assessment for the Dementing Elderly (PADE) (Villanueva et al., 2003) | r = 0.30 - 0.42 with CMAI |
| Pain Assessment Checklist for Seniors with Limited Ability to Communicate) (PACSLAC) (Fuchs-Lacelle et al., 2004) | r=0.34 with Doloplus 2 |
| Behavioral Pain Scale (BPS) (Payen et al., 2001) | Not available |
| Doloplus-2 (Lefebvre-Chapiro, 2001) | r = 0.29-0.38 with PACSLAC |
| NOPPAIN (Non-Communicative Patient's Assessment Instrument) (Snow et al., 2004) | Not Available |
| NVPS: Nonverbal Pain Scale (Odhner et al., 2003b) | Not Available |
| Communication Effectiveness Index (CETI) (Lomas et al., 1989) | r = 0.61with Western Aphasia Battery |
| Boston Diagnostic Aphasia Examination (BDAE) (Goodglass and Kaplan, 1982) | Not Available |
| The Frenchay Aphasia Screening Test (Enderby et al., 1987) | r = 0.73 with Functional Communication Profile |
| Porch Index of Communicative Abilities (PICA) (Porch, 1971) | r = 0.63 with CET, r = - 7.49 with Western Aphasic Battery WAB |
| Communication Activities of Daily Living -2 (CADL-2) (Holland, 1998) | Not Available |
| ASHA Functional Assessment of Communication Skills (ASHA FACS) (Frattali et al 1995) | r = 0.63 with ADAS Cog, r = 0.72 with MMSE |
| Kovacev Non-Verbal Expression Check List (KNVECL) (Kovacev, 2008) | Not Available |
| ADD (Assessment of Discomfort in Dementia) (Kovach et al., 1999) | Not Available |
| Lille Communication Test (LCT) (Rousseaux, 2001) | Not Available |
| The Holden Communication Scale (HCS) (Holden, 1995) | $r = -0.67$ with MMSE |
| Quality of Interaction schedule (QUIS) (Dean et al., 1993) | Not Available |
| The Assessment of Communication and Interaction Skills (ACIS) (Forsyth et al., 1999) | Not Available |
| Core Linguistic Battery (Bayles et al., 1992) | Not Available |
| Communication Problems Scale (Powell et al., 1995) | Not Available |
| Emotion Recognition Test (ERT) (Shimokawa et al 2000) | $r = 0.53$ ($p < 0.004$) between Emotion Recognition Score and Interpersonal Behaviour Checklist |
| Multidimensional Observation Scale for Elderly Subjects (MOSES) (Helmes et al., 1987) | $r = 0.49$ ($p < 0.001$) between Moses Depression Anxiety and Zung Depression Status Inventory |
| Posture Knowledge Test (Mozaz et al., 2002) | Not Available |
| Social Competence Scale (Westermeyer and Harrow, 1986) | Not Available |
| The Functional Linguistic Communication (FLCI) Inventory (Bayles., et al 1994) | Not Available |
| Comprehensive Assessment of Spoken Language (CASL) (Carrow-Woolfolk, 1999) | $r = 0.74$ with Peabody Picture Vocabulary Test-Third Edition |
| Facial Action Coding System (FACS) (Ekman et al., 1978) | Between pain scale and proxy pain report) $n=26$ AU (frequency), $r=0.62$; AU (intensity) $r=0.73$; |
| Verbal Fluency Benton (FAS) (Benton et al., 1976) | $r = 0.25$ with Vocabulary scores (WAIS-R) |
| Interpersonal Communication Inventory (Bienvenu, 1971) | Not Available |
| The Adult Behavior Questionnaire (ABQ) (Magai et al., 1996) | $r = 0.36$ between facial expressions and ABQ caregiver report interest item, $r = 0.56$ between facial expressions and joy item on ABQ caregiver report. $R = 0.61$ between ABQ-F anger item and facial expressions |

**Table 5.**
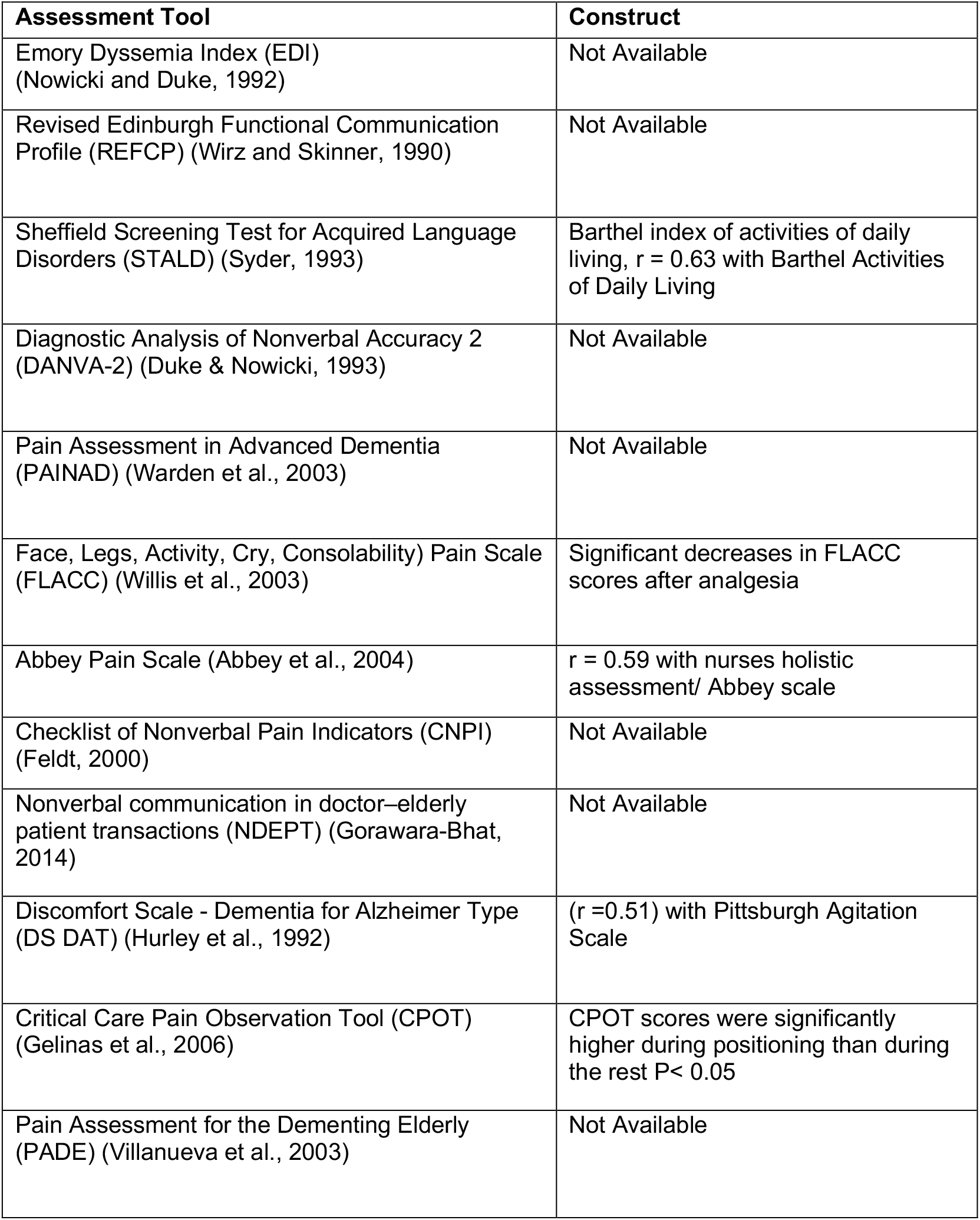
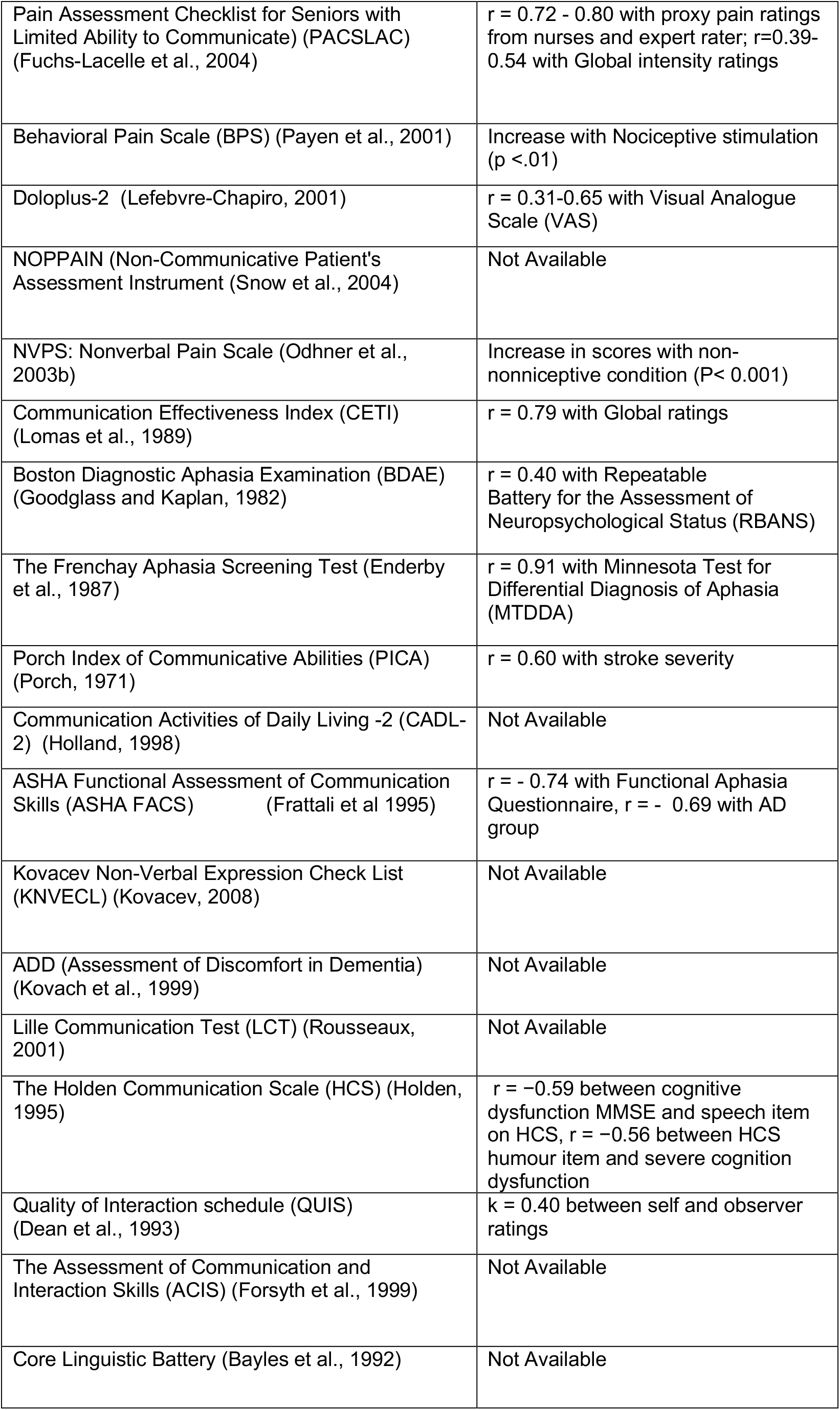
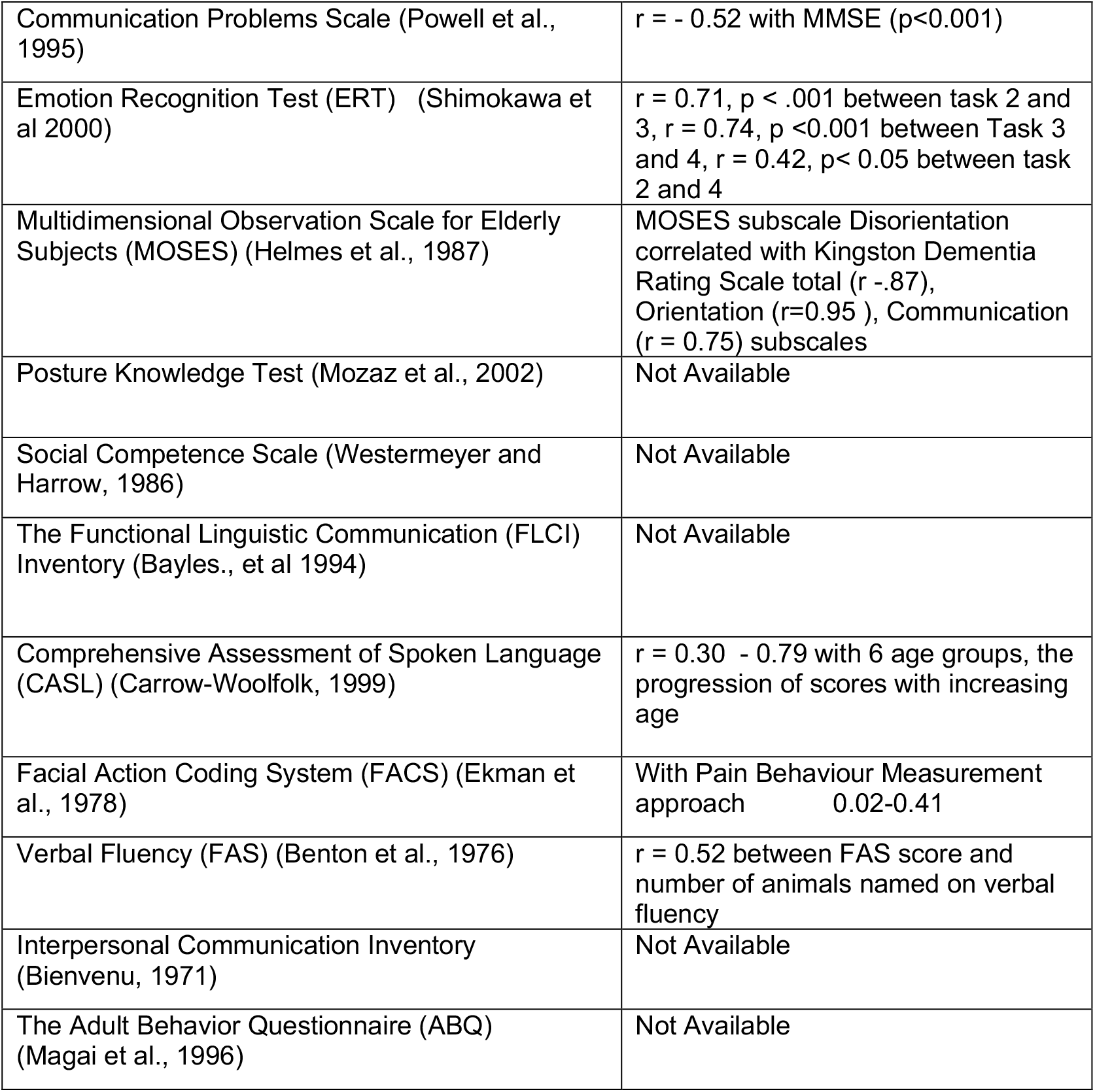
Construct Validity of Tools Identified.

**Table 6.**
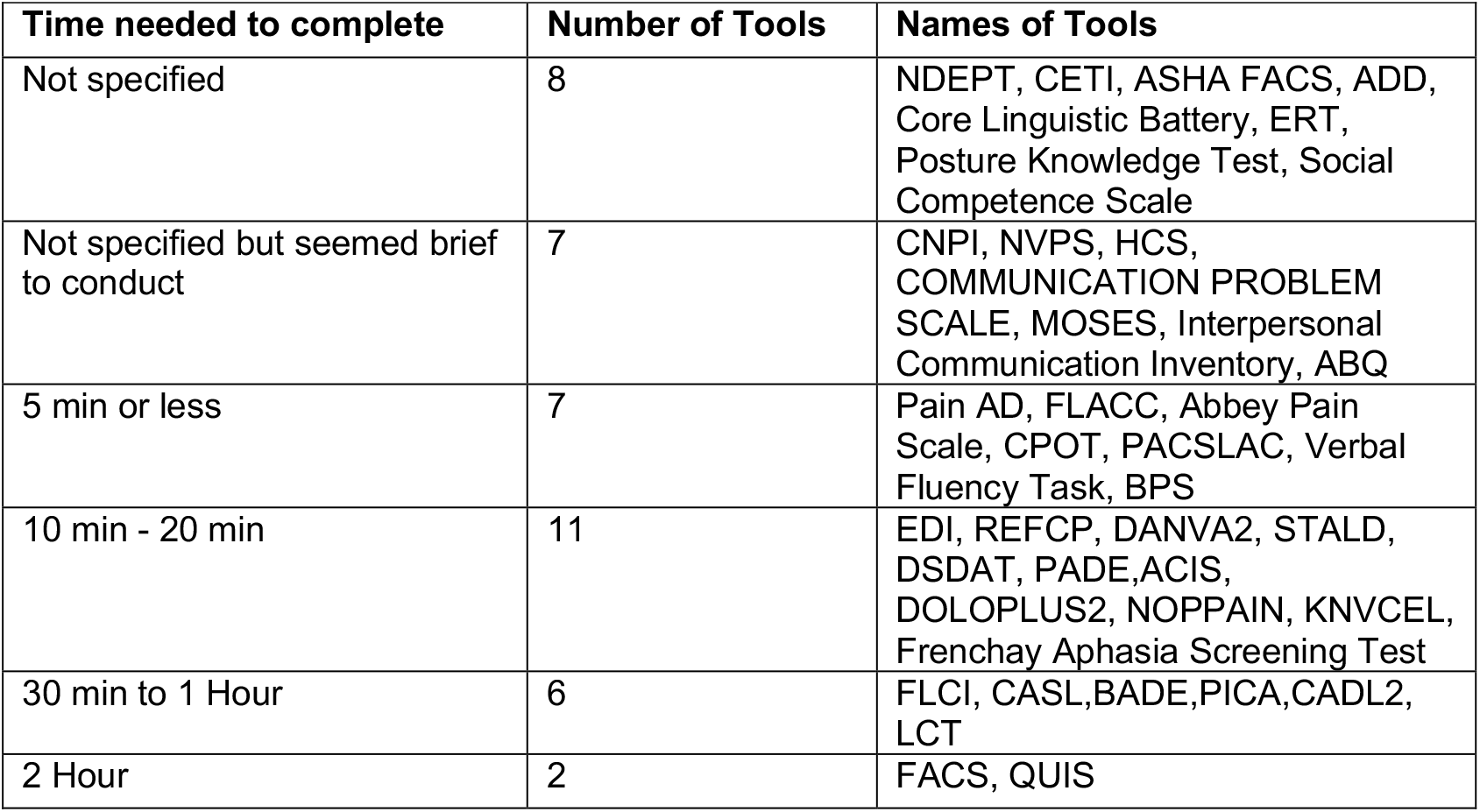
Time needed to complete the assessment.

| **Time needed to complete** | **Number of Tools** | **Names of Tools** |
| --- | --- | --- |
| Not specified | 8 | NDEPT, CETI, ASHA FACS, ADD, Core Linguistic Battery, ERT, Posture Knowledge Test, Social Competence Scale |
| Not specified but seemed brief to conduct | 7 | CNPI, NVPS, HCS, COMMUNICATION PROBLEM SCALE, MOSES, Interpersonal Communication Inventory, ABQ |
| 5 min or less | 7 | Pain AD, FLACC, Abbey Pain Scale, CPOT, PACSLAC, Verbal Fluency Task, BPS |
| 10 min - 20 min | 11 | EDI, REFCP, DANVA2, STALD, DSDAT, PADE, ACIS, DOLOPLUS2, NOPPAIN, KNVCEL, Frenchay Aphasia Screening Test |
| 30 min to 1 Hour | 6 | FLCI, CASL, BADE, PICA, CADL2, LCT |
| 2 Hour | 2 | FACS, QUIS |

**Table 7.**
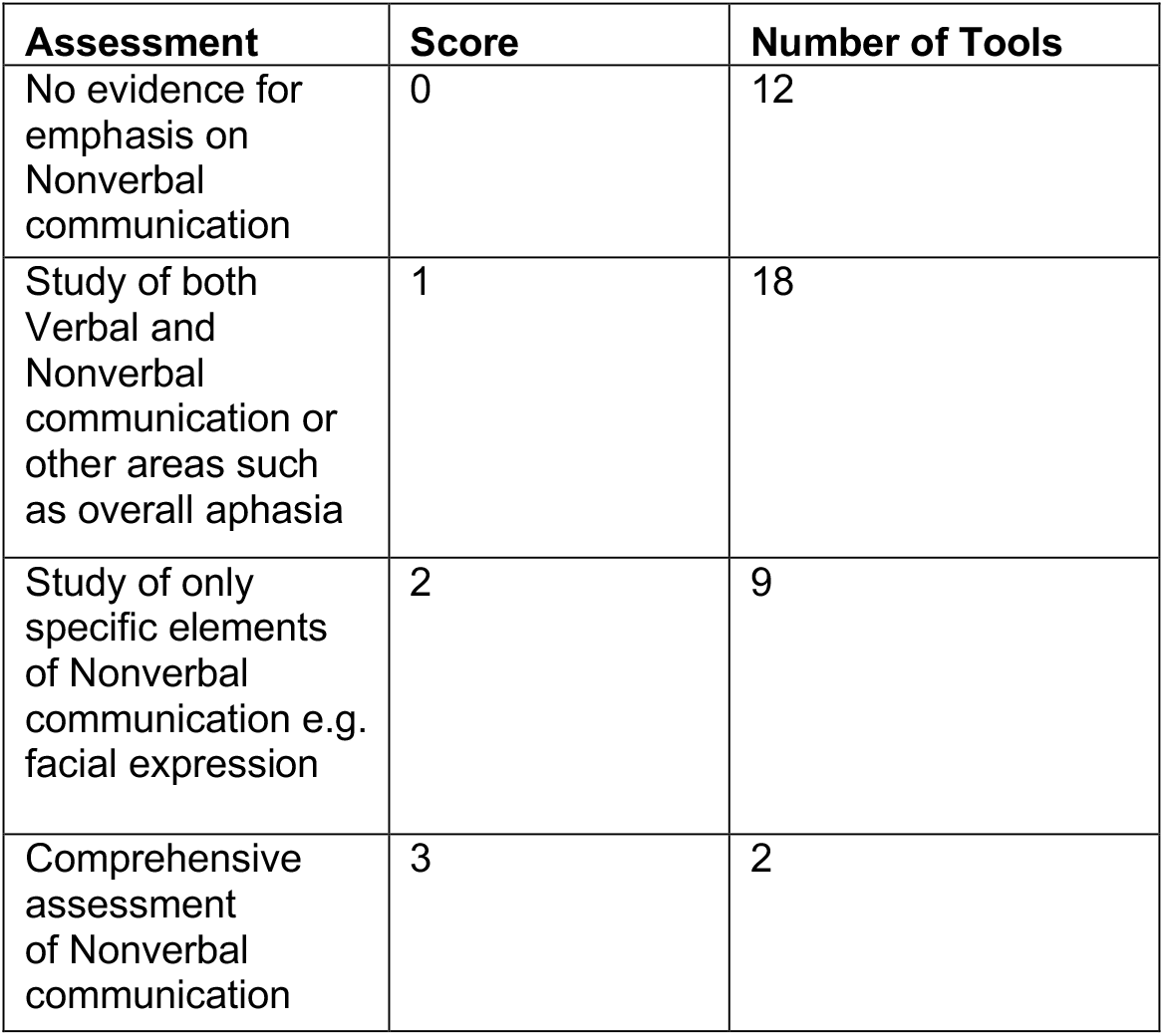
Suitability scores for tools: (Coverage of Nonverbal Communication)

| **Assessment** | **Score** | **Number of Tools** |
| --- | --- | --- |
| No evidence for emphasis on Nonverbal communication | 0 | 12 |
| Study of both Verbal and Nonverbal communication or other areas such as overall aphasia | 1 | 18 |
| Study of only specific elements of Nonverbal communication e.g. facial expression | 2 | 9 |
| Comprehensive assessment of Nonverbal communication | 3 | 2 |

### Content validity

The majority of the tools did not focus on nonverbal communication alone but in conjunction with other aspects of communication including functional and verbal communication. A review of the content validity was undertaken in relation to the ability to assess nonverbal communication for all identified tools. Using the criteria based on a methodology used in previous literature review [51] and Streiner and Norman’s requirements Streiner [9] for health measurement scales, the majority of the tools covered important items/dimensions to a moderate extent only; and three tools had essential items and domains of nonverbal communication function. Eleven tools did not appear to cover the important items/dimensions.

All of the measures outlined were developed for use within a diverse range of settings and purposes. The majority of the measures were adapted from measures originally developed for a different patient group, from young children (DANVA, EDI, FLACC, Doloplus-2) to PlwD (PAINAD) and Abbey Pain scale. Furthermore, some tools were developed for specific users (QUIS for professionals; NOPPAIN, for nursing assistants), another for research purposes (DS-DAT) and as decision support tools (the ADD Protocol). During the review process of the communication tools, five distinct areas emerged, in which tools could be grouped together. These tools were categorised into distinct categories, as listed below. See Appendix A and B for detail information on psychometric and feasibility qualities.

### Categories of Communication Tool

- Nonverbal communication
- Stroke and Aphasia testing verbal communication
- Nonverbal Communication tools within the context of Pain
- Functional communication and overall communication function
- Tools assessing multidimensional function

**Figure 4.**
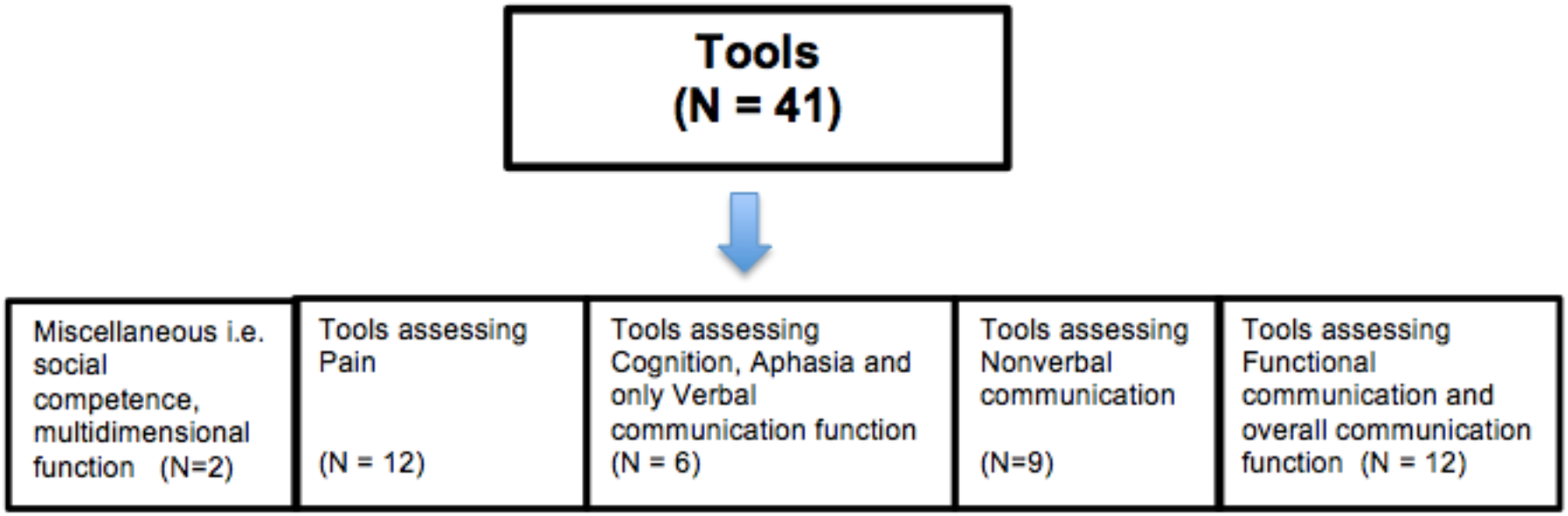
Identified Communication Tools.

### Tools assessing Nonverbal Communication

In total, only nine of the 41 tools specifically examined nonverbal communication; a substantial proportion of these tools did not exhibit a comprehensive assessment of nonverbal communication function and specific indices of nonverbal communication indices. These tools often only assessed specific aspects of nonverbal communication, e.g. facial expressions via DANVA-2 [14] and the emotion recognition task [41]. Other key examples included Nonverbal communication in Doctor–Elderly Patient Transactions (NDEPT) [19] which reflected upon interactions between clinicians and their patients. One tool which reflected a detailed study of nonverbal cues was the Emory Dyssemia Index (EDI) [52].

### Assessment of Nonverbal Communication within the context of Pain

A substantial proportion of tools investigating elements of nonverbal communication originated from pain related scales (n=12), including the FLACC (Face, Legs, Activity, Cry, Consolability) Pain Scale [16], Pain Assessment in Advanced Dementia [15] and DS-DAT (Discomfort Scale − Dementia for Alzheimer Type) [20]. These nonverbal pain scales evaluated expressions related to pain (i.e., negative or positive facial expression, grimaces, furrowed brow, smile and frown) in comparison to assessing overall communication function or specific nonverbal cues.

### Assessment of Functional Communication and Overall Communication Function

Other tools covered some elements of nonverbal communication which derived from measures examining functional communication (n= 12) where both verbal and nonverbal communication modalities are examined to assess the efficacy of the message being conveyed during daily communication interactions. Two examples of this were the Revised Edinburgh Functional Communication Profile (REFCP) [12] and the ASHA Functional Assessment of Communication Skills (ASHA FACS) [11]. Other tools which formed a multidimensional assessment of function or social competence were also reviewed in the process, these were grouped under the miscellaneous category.

### Stroke and Aphasia testing verbal communication

Some measures that were highlighted in the current review were derived from tools examining acquired language disorders such as aphasia, these exhibited tasks items assessing cognitive functioning and verbal reasoning, such as Boston Diagnostic Aphasia Examination (BDAE) [29] and other brief tests e.g. FAS Verbal Fluency. In addition, some tools only focussed purely on verbal aspects, for instance, the Sheffield Screening Test for Acquired Language Disorders [13] that has been widely used in studies with people living with dementia to test receptive and expressive language function via verbal comprehension for single words and word-finding ability. In total, six out of forty-one tools came across as an assessment of aphasia, testing verbal functioning in people with or without dementia. These assessments were reviewed alongside other scales, with the aim to seek tools examining nonverbal communication in PlwAD.

### Overall Psychometric Quality Evaluation

Several tools scored high on psychometric properties, for instance, assessment scales for pain including Face, Legs, Activity, Cry, Consolability) Pain Scale (FLACC) [16], Nonverbal Pain Scale (NVPS) [27], Behavioural Pain Scale (BPS) [24] and Discomfort Scale - Dementia for Alzheimer Type (DS DAT) [20]. Other tools scoring good included tests for aphasia and spoken language; Emory Dyssemia Index, ASHA Functional Assessment of Communication Skills (ASHA FACS) [11], Communication Effectiveness Index [28], Frenchay Aphasia Screening Test [30] and Porch Index of Communicative Ability [31]. One tool specifically focussed on nonverbal communication was the Facial Action Coding System for measuring facial expression which also managed to score towards higher end.

In contrast, some tools failed to score high on psychometric qualities. These were Core Linguistic battery [39], The Functional Linguistic Communication [45], Revised Edinburgh Functional Communication Profile [12], Social Competence Scale [44], Checklist of Nonverbal Pain Indicator (CNPI) [18], NOPPAIN (Non-Communicative Patient’s Assessment Instrument [26], Lille Communication Test (LCT) [35] and Posture Knowledge Test [43].

### Feasibility and Time Taken to Complete

Evidence available for feasibility and clinical utility of tools was scarce and the criteria for scoring and interpreting scores were often not readily available. Twelve tools that scored zero and appeared complex to conduct included the NDEPT, PACSLAC, NVPS, BDAE, The Frenchay Aphasia Screening Test, PICA, CADL-2, LCT, Core Linguistic Battery, Posture Knowledge Test, CASL, FACS. Several tools that were manageable in a set format comprised of STALD, DANVA-2, PAINDAD, Doloplus-2, NOPPAIN, KNVECL, ADD, QUIS, ERT, Verbal Fluency FAS and FLCI. However 18 tools scored two (i.e. the if tool was short, manageable with instructions, scoring interpretation), this list of tools included the EDI, REFCP, FLACC, Abbey Pain Scale, CNPI, DS DAT, CPOT, PADE, BPS, CET, ASHA-FACS, HCS, ACIS, Communication Problem Scale, MOSES, Social Competence Scale, Interpersonal Communication Inventory and ABQ.

Time taken to complete was also recorded, whenever possible. Information in relation to the time needed to complete an assessment was not specified for eight tools. Some tools did not specify the time requirement; however, these tools were present in a set format that seemed brief to conduct. Only two of the tools were suggested to have duration of two hours.

### Suitability to Assess Nonverbal Communication

Suitability assessed coverage of nonverbal communication within tools; 12 of the 41 tools did not offer an assessment of nonverbal communication, for instance, these focussed on assessing verbal communication purely, namely Sheffield Screening Test for Acquired Language Disorders [13] and Comprehensive Assessment of Spoken Language (CASL) [46]. These tools scored zero for assessing nonverbal communication. Several other tools covered elements of both verbal and nonverbal communication (19/41), these included tools with the score of one for the ability to assess nonverbal cues, examples of such tools integrated tools like the Revised Edinburgh Functional Communication Profile (REFCP) [12] and some of the pain assessing scales, namely Pain Assessment for the Dementing Elderly (PADE) [22]. However other tools encompassed the study of a specific nonverbal cue, e.g. facial expressions through the Facial Action Coding System [53], and these tools received a score of two for assessing nonverbal communication (13/41). Only three tools were identified to provide a comprehensive study of different nonverbal communication cues. The Emory Dyssemia Index (EDI) examined impairment in nonverbal communication through a number of nonverbal communication cues including gesture, gaze, facial expressions and space and touch using.

### Findings of Extended Narrative review

The extended narrative review from 2017 to October 2019 identified 272 papers, of which 49 were retained in the search following the titles being screened. Of these 20 papers focussed on the assessment of pain, four focussed on overall communication; 18 focussed on emotion recognition in addition to seven papers that focussed on nonverbal communication (including gestures, touch and overall nonverbal communication). Two tools were found to examine elements nonverbal communication. The Threadgold Communication tool (TCT) [4, 54] focussed on difficulties in the number of communication cues, including speaking, vocal sounds, singing, smell, gestures, interactive touch and posture. The TCT consists of 14-item questionnaire, and each item is graded from zero to four scale, with zero indicating no evidence to four indicating frequent evidence of communication.

Evidence presented on psychometrics properties of this tool reflect a satisfactory level for internal consistency and a high test-retest reliability (r = 0.76). The corrected item-total correlation ranged between 0.50 and 0.87. Despite the modest sample size (n = 51), the TCT is a reliable and valid instrument, suitable for measuring communication among people with moderate to severe dementia pre- and post-intervention. The second tool identified to assess elements of nonverbal communication was the Scenario Test (ST) [55], a battery test which examines the ability to convey a message verbally and/or nonverbally, in every-day life scenarios. The test was designed for persons with severe to moderate limitations of verbal communication for people with severe aphasia testing an individual’s ability to communicate in a set scenario. Despite limited evidence of use in clinical practice and research, there is some evidence for validity and reliability of the tool, high internal consistency (α =0.96) and retest reliability (ICC: 0.98) was present. Concurrent validity for ST is reported as good with other tools in aphasia (0.85 − 0.79).

## Discussion

### Summary of Findings

The objective of the literature review was to identify tools available to assess nonverbal communication that could be potentially used for assessing nonverbal communication in people living with AD. Forty-one tools were identified through the scoping review (1946 - 2017), these tools originated and focussed on different forms of communication and were used with various populations and settings.

The review revealed a limited number of tools available that focussed purely on investigating nonverbal communication (n = 9). Some of the tools developed were specifically for assessing nonverbal communication, this included: EDI, DANVA-2, NDEPT, KNVECL, ERT, Posture Knowledge Test, FACS and ABQ. Pain and functional communication were the two most common focuses for these tools. These tools utilised different methodological approaches to assess nonverbal communication, most were observational and used test batteries and proxy questionnaires. The findings of the scoping review were further confirmed by the recent narrative literature search process reported in this paper, which also indicates towards a lack of nonverbal research tools present within the current literature. Limited number scales provided a detailed report of nonverbal communication function,

### Findings in Context of Research

The scoping review aimed to identify nonverbal communication tools that can be employed to examine nonverbal communication in PlwD. Limited literature was available on nonverbal communication in dementia, it is likely that this is partly due to the lack of reliable tools. Furthermore, to date there are no research review papers present that focus on ways of assessing nonverbal communication cues, particularly in dementia. As a whole there is limited evidence to support the feasibility and validity of using these tools in PlwD. This is due to a variety of reasons including the method of data collection, duration and the presence of cognitive impairment in this population. Data on feasibility was insufficient across majority of tools, commonly due to limited information provided on feasibility within the original studies. Variation in settings and population may have an influence on psychometric findings of individual measures, for example in an acute setting; ratings might be affected by having limited knowledge of the participants and disease severity.

Unlike verbal language, the innate nature of nonverbal communication is continuous with multiple channels, contexts and meanings, thus making this a challenging form of communication to assess and define. For instance, aspects of nonverbal communication, such as changes in voice tone and body language could take place without a person’s explicit awareness [56], and often varying in their frequency and intensity. Verbal and nonverbal communication cues typically occur simultaneously and are interpreted together during face-to-face interactions [57, 58]. These factors add to the complexity to the evaluation of nonverbal cues and their associated context, meaning and relevance.

Very few tools appeared to have any strong theoretical underpinning which supported the development of the tool and there was variation in research methodologies and calculations for determining psychometric properties. Therefore, a direct comparison across the tools should be interpreted with caution. Overall, the current review supports the previous literature indicating a clear gap in research in relation to nonverbal communication within the previous literature [59]. Further research is warranted within this field to fill in the gaps and to identify appropriate research methodologies for assessing nonverbal communication across various populations and settings.

### Limitations and Strength

The review was kept broad in terms of study population and setting, and this represents a limitation of this review. The studies identified in this review have considerable heterogeneity, in terms of design (retrospective vs. prospective), method (observational tools vs. batteries), research population (dementia, stroke, aphasia, different levels of cognitive impairment and different settings), making their data difficult to compare.

In addition, assumptions about the psychometric quality of the tools were based on the information provided and methodological details were often omitted from the report. Further elements that make the studies difficult to compare include differences in format/structure and scoring method. For instance, DOLOPLUS2, PADE, BDAE, REFCP ERT, and FACS are very different in these respects. A set of criteria was used to provide an indication of psychometric strengths, which allowed the review to be more objective. However, the quality scores should always be interpreted with caution since the use of some criteria involved a subjective element derived from the reviewer’s expertise. The current literature search was kept broad to capture the presence of nonverbal tools and did not specifically target literature reporting on the feasibility for the tools, because of this, the evidence may be more limited on these properties than it might have been otherwise. The findings of this review help to provide insight into the different research approaches that can be used to assess communication function, approaches which could be developed for use in the dementia population.

### Implications for Research and Practice

There are various research and practice implications for the findings of this review. This review could be expanded to include distinct categories of nonverbal cues in the literature search process to ensure broader and fuller coverage of nonverbal research literature. There were forty-one tools identified through the scoping review which included elements of nonverbal cues. However, we cannot at present recommend any particular tool for use in any clinical practice, due to the lack of comprehensive evidence on the reliability, validity, feasibility or clinical utility of any one particular tool. Further research should be conducted on the psychometric properties of tools and in clinical practice to assess feasibility, clinical utility and guidance on use of the tools. These are necessary albeit not sufficient pre-requisites for actual use in routine clinical practice.

Furthermore, there is a need to evaluate the nonverbal tools for people living with dementia across various clinical settings, using rigorous methods and larger sample sizes. These studies need to ensure that the definitions and stages of cognitive impairment are clearly defined, to enable comparison of nonverbal communication function across disease severity. Whilst there is a need for rigorous research, assessing the accuracy of tools can be difficult due to the various methodological research approaches being used and there being no clear gold standard approach [60]. Thus, researchers need to either envision new assessment instruments on the basis of different conceptual foundations or concentrate on extending the psychometric evidence for some of the existing tools. It should also be noted that whilst some tools were identified to have high reliability in research this may not be transferable to routine clinical practice because many of these tools (e.g., particularly pain or EDI) were not being employed as intended.

Literature indicates that nonverbal communication plays an important role in conveying emotional and relational information during clinical interactions [61-63]. This review adds to our understanding of the availability of nonverbal communication measures that could be adapted for various settings, Whilst these tools may be adaptable to different setting their applicability to the real life setting in care homes, for the dementia population this remains unknown [64]. Larger sample size studies will be needed to evaluate feasibility and practical use of identified tools across different populations and settings. Finally, a tool must be applicable to “the condition, setting and individuals participating in an interaction” [65]. This understanding also highlights the importance of context features and emphasizes the need to adopt a person-centred approach, to the extent to which a measure captures both the content of the health care user’s views and the form or ways in which their views are expressed [66].

Several communication tools assessing nonverbal behaviour (such as FACS, NDEPT, KNVS) are observational tools utilising real time coding and video analysis, which can be intrusive and resource intensive [67]. An assessment of nonverbal function could be conducted using a proxy assessment, using the EDI scale. Such tools can be further evaluated for validity, reliability and feasibility on a wider scale to match the needs of individuals with dementia. This tool should be studied further to assess its feasibility for use with people living with dementia, matching the possible urgency for such a tool, particularly to improve the well-being of people living in the severe stages of dementia who are not able to communicate their needs. In summary, there a limited number of nonverbal communication tools present currently available. The primary objective of this review was to identify tools for evaluation of nonverbal communication function with a view of utilizing the tool pre- and post-intervention to focus on nonverbal cues as essential means of communication for PlwD in care homes. In line with this, future work is needed to develop a tool specifically focussed on nonverbal communication function.

## Conclusion

This review highlights the current evidence base of nonverbal communication assessment tools and provides an insight regarding the gaps in our understanding. Forty-one tools were identified through the literature search process. These tools assessed various communication channels embracing verbal, nonverbal and functional, communication means; originating from various settings and populations, for instance, those assessing cognition and verbal language difficulties secondary to stroke, aphasia and nonverbal cues associated with pain. A number of tools presented psychometrics qualities; only nine of the forty-one tools specifically focussed on nonverbal communication and most of these tools did not provide a comprehensive assessment of nonverbal communication function. It is therefore challenging to recommend one particular tool for use in any clinical or care setting, due to the mismatch of suitability, reliability, validity, and feasibility of any one particular tool. However, namely two tools including the EDI scale and the TCT indicated coverage of nonverbal communication across spectrum of different nonverbal cues. The EDI scale indicated a proxy assessment of impairment across different nonverbal communication skills including paralanguage, gestures facial expressions chronemics and use of space and touch with good test re-test reliability and internal consistency, however, requires further testing to validate it’s use in dementia. The EDI scale provides extensive study of nonverbal cues, with the advantage of requiring short time and brief minimum instructions and burden for caregivers to complete, the higher the score, the worse the impairment. The Threadgold Communication Tool (TCT) focussed on difficulties in the number of communication cues. TCT provided satisfactory level for internal consistency and a high test-retest reliability for use in dementia (albeit small sample size). Compared to the TCT, the EDI scale covered nonverbal communication in a greater depth. Further research is required to evaluate psychometric properties and in clinical or care practice to assess feasibility, clinical utility and guidance for application in dementia for both EDI and TCT.

## Data Availability

The authors confirm that the data supporting the findings of this study are available within the article [and/or] its supplementary materials.

## Appendix A Identified tools and Psychometric Properties

| Assessment Tool | Origin of Item - Population | Number of Participants | Content | Construct | Criterion | Homogeneity (internal consistency) | Inter-rater Reliability | Re-test Reliability | Coverage of nonverbal communication | Sum for Psychometric properties |
| --- | --- | --- | --- | --- | --- | --- | --- | --- | --- | --- |
| **Selected Tools** |  |  |  |  |  |  |  |  |  |  |
| Emory Dyssemia Index (EDI) (Nowicki and Duke, 1992) | 0 | 2 | 2 | 0 | 2 | 2 | 0 | 2 | 3 | 8 |
| Revised Edinburgh Functional Communication Profile (REFCP) (Wirz SL, Skinner C, 1990) | 0 | 1 | 1 | 0 | 0 | 0 | 1 | 0 | 1 | 2 |
| Sheffield Screening Test for Acquired Language Disorders (Syder, 1993) | 1 | 2 | 0 | 2 | 2 | 0 | 0 | 0 | 0 | 4 |

| Non-Selected Tools |  |  |  |  |  |  |  |  |  |  |
| --- | --- | --- | --- | --- | --- | --- | --- | --- | --- | --- |
| DANVA-2<br>(Diagnostic Analysis of Nonverbal Accuracy 2)<br>(Duke & Nowicki, 1993) | 0 | 2 | 1 | 0 | 1 | 2 | 0 | 2 | 2 | 6 |
| Pain Assessment in Advanced Dementia (PAINAD)<br>(Warden et al., 2003) | 1 | 0 | 1 | 0 | 2 | 1 | 2 | 0 | 2 | 6 |
| FLACC (Face, Legs, Activity, Cry, Consolability)<br>Pain Scale (Willis et al., 2003) | 1 | 1 | 1 | 2 | 2 | 2 | 2 | 0 | 2 | 9 |
| Abbey Pain Scale (Abbey et al., 2004) | 1 | 2 | 1 | 0 | 1 | 2 | 0 | 0 | 2 | 4 |
| CNPI (Checklist of Nonverbal Pain Indicators)<br>(Feldt, 2000) | 1 | 1 | 1 | 0 | 1 | 1 | 2 | 0 | 1 | 5 |
| Nonverbal communication in doctor-elderly patient transactions (NDEPT) (Gorawara-Bhat, 2014) | 1 | 1 | 2 | 0 | 0 | 0 | 1 | 0 | 2 | 3 |
| Discomfort Scale - Dementia for Alzheimer Type (DS DAT) (Hurley et al., 1992) | 2 | 2 | 1 | 2 | 1 | 1 | 2 | 1 | 2 | 8 |
| Critical Care Pain Observation Tool (CPOT) (Gelinas et al., 2006) | 0 | 0 | 1 | 1 | 1 | 2 | 2 | 0 | 1 | 7 |
| Pain Assessment for the Dementing Elderly (PADE) (Villanueva et al., 2003) | 2 | 1 | 1 | 0 | 1 | 1 | 2 | 1 | 1 | 6 |
| PACSLAC (Pain Assessment Checklist for Seniors with Limited Ability to Communicate) (Fuchs-Lacelle et al., 2004) | 2 | 2 | 1 | 2 | 1 | 2 | 0 | 0 | 1 | 6 |
| Behavioral Pain Scale (BPS) (Payen et al., 2001) | 0 | 1 | 1 | 2 | 0 | 2 | 1 | 0 | 1 | 6 |
| Doloplus-2 (Lefebvre-Chapiro, 2001) | 1 | 2 | 0 | 0 | 1 | 2 | 1 | 0 | 0 | 4 |
| NOPPAIN (Non-Communicative Patient's Assessment Instrument (Snow et al., 2004) | 1 | 0 | 1 | 0 | 0 | 0 | 0 | 0 | 1 | 1 |
| NVPS: Nonverbal Pain Scale (Odhner et al., 2003b) | 0 | 0 | 1 | 2 | 1 | 1 | 2 | 1 | 1 | 8 |
| Communication Effectiveness Index (CETI) (Lomas et al., 1989) | 0 | 0 | 0 | 2 | 1 | 2 | 1 | 2 | 0 | 8 |
| Boston Diagnostic Aphasia Examination (BDAE) (Goodglass and Kaplan, 1982) | 1 | 2 | 1 | 1 | 1 | 1 | 0 | 0 | 1 | 4 |
| The Frenchay Aphasia Screening Test (Enderby et al., 1987) | 0 | 0 | 0 | 2 | 2 | 0 | 2 | 2 | 0 | 8 |
| Porch Index of Communicative Abilities (PICA) (Porch, 1971) | 1 | 1 | 1 | 1 | 2 | 0 | 0 | 2 | 1 | 6 |
| Communication Activities of Daily Living -2 (CADL-2) (Holland, 1998) | 1 | 2 | 1 | 0 | 0 | 2 | 0 | 2 | 1 | 5 |
| ASHA Functional Assessment of Communication Skills (ASHA FACS) (Frattali et al 1995) | 2 | 2 | 1 | 2 | 2 | 2 | 2 | 2 | 1 | 11 |
| Kovacev Non-Verbal Expression Check List (KNVECL) (Kovacev, 2008) | 2 | 2 | 2 | 0 | 0 | 2 | 0 | 0 | 3 | 4 |
| ADD (Assessment of Discomfort in Dementia) (Kovach et al., 1999) | 2 | 2 | 1 | 0 | 0 | 0 | 2 | 0 | 1 | 3 |
| Lille Communication Test (LCT) (Rousseaux, 2001) | 0 | 0 | 1 | 0 | 0 | 0 | 2 | 0 | 1 | 3 |
| The Holden Communication Scale (Holden, 1995) | 2 | 2 | 0 | 1 | 1 | 2 | 0 | 1 | 0 | 5 |
| Quality of Interaction schedule (Dean et al., 1993) | 2 | 2 | 0 | 1 | 0 | 0 | 1 | 0 | 0 | 2 |
| The Assessment of Communication and Interaction Skills (ACIS) (Forsyth et al., 1999) | 1 | 2 | 0 | 2 | 1 | 0 | 2 | 0 | 1 | 5 |
| Core Linguistic Battery (Bayles et al., 1992) | 1 | 2 | 0 | 0 | 0 | 0 | 0 | 0 | 0 | 0 |
| Communication Problems Scale (Powell et al., 1995) | 2 | 2 | 1 | 1 | 0 | 2 | 0 | 0 | 0 | 4 |
| Emotion Recognition Test (Shimokawa et al 2000) | 2 | 0 | 1 | 1 | 1 | 1 | 0 | 0 | 2 | 4 |
| Multidimensional Observation Scale for Elderly Subjects (MOSES) (Helmes et al., 1987) | 1 | 2 | 0 | 1 | 1 | 2 | 2 | 0 | 0 | 6 |
| Posture Knowledge Test (Mozaz et al., 2002) | 2 | 1 | 0 | 0 | 0 | 0 | 0 | 0 | 2 | 0 |
| Social Competence Scale (Westermeyer and Harrow, 1986, Zigier-Phillips 1961) | 1 | 0 | 0 | 0 | 0 | 0 | 0 | 0 | 0 | 0 |
| The Functional Linguistic Communication (Bayles., et al 1994) | 2 | 2 | 1 | 0 | 0 | 0 | 0 | 0 | 1 | 1 |
| Comprehensive Assessment of Spoken Language (CASL) (Carrow-Woolfolk, 1999) | 0 | 2 | 0 | 2 | 2 | 2 | 0 | 2 | 0 | 8 |
| Facial Action<br>Coding System<br>(Ekman et al.,<br>1978) | 1 | 2 | 2 | 1 | 2 | 0 | 2 | 0 | 1 | 7 |
| Verbal Fluency<br>Benton (Benton<br>et al., 1976) | 2 | 2 | 0 | 1 | 1 | 0 | 0 | 1 | 1 | 3 |
| Interpersonal<br>Communication<br>Inventory<br>(Bienvenu,<br>1971) | 0 | 2 | 0 | 0 | 0 | 2 | 1 | 2 | 0 | 5 |
| The Adult<br>Behaviour<br>Questionnaire<br>(ABQ) (Magai et<br>al., 1996) | 2 | 2 | 2 | 0 | 1 | 1 | 0 | 0 | 2 | 4 |

## Appendix B Communication Tools

| Assessment Tool | Feasibility Score (Manageable or complex to conduct) |
| --- | --- |
| Emory Dyssemia Index (EDI) | 2 |
| Revised Edinburgh Functional Communication Profile (REFCP) | 2 |
| Diagnostic Analysis of Nonverbal Accuracy 2 (DANVA-2) | 1 |
| Sheffield Screening Test for Acquired Language Disorders (STALD) | 1 |
| Pain Assessment in Advanced Dementia (PAINAD) | 1 |
| FLACC (Face, Legs, Activity, Cry, Consolability) Pain Scale | 2 |
| Abbey Pain Scale | 2 |
| CNPI (Checklist of Nonverbal Pain Indicators) | 2 |
| Nonverbal communication in doctor–elderly patient transactions (NDEPT) | 0 |
| Discomfort Scale - Dementia for Alzheimer Type (DS DAT) | 2 |
| Critical- Care Pain Observation Tool (CPOT) | 2 |
| Pain Assessment for the Dementing Elderly (PADE) | 2 |
| Pain Assessment Checklist for Seniors with Limited Ability to Communicate (PACSLAC) | 0 |
| Behavioral Pain Scale (BPS) | 2 |
| Doloplus-2 | 0 |
| NOPPAIN (Non-Communicative Patient's Assessment Instrument) | 1 |
| NVPS: Nonverbal Pain Scale | 0 |
| Communication Effectiveness Index (CETI) | 2 |
| Boston Diagnostic Aphasia Examination (BDAE) | 0 |
| The Frenchay Aphasia Screening Test (FAST) | 0 |
| The Functional Linguistic Communication Inventory. | 1 |
| Porch Index of Communicative Abilities (PICA) | 0 |
| Communication Activities of Daily Living -2 (CADL-2) | 0 |
| ASHA Functional Assessment of Communication Skills (ASHA FACS) | 2 |
| Kovacev Nonverbal Expression Check List (KNVECL) | 1 |
| ADD (Assessment of Discomfort in Dementia) | 1 |
| Lille Communication Test (LCT) | 0 |
| The Holden Communication Scale | 2 |
| Quality of Interaction schedule (QUIS) | 1 |
| The Assessment of Communication and Interaction Skills (ACIS) | 2 |
| Core Linguistic Battery | 0 |
| Communication Problems Scale | 2 |
| Emotion Recognition Test | 1 |
| Multidimensional Observation Scale for Elderly Subjects | 2 |
| Posture Knowledge Test | 0 |
| Social Competence Scale | 2 |
| Facial Action Coding System | 0 |
| Interpersonal Communication Inventory | 2 |
| Verbal Fluency | 1 |
| Comprehensive Assessment of Spoken Language (CASL) | 0 |
| The Adult Behavior Questionnaire | 2 |

